# Periosteal pressure sensitivity–guided non-pharmacological intervention lowers cardiovascular event rates after five years in ischemic heart disease: Evidence from a randomized controlled trial

**DOI:** 10.64898/2026.05.27.26354261

**Authors:** S. Ballegaard, F. Gyntelberg, A. S. Afzal, A. Hjalmarson, J. Faber

**Author notes:** Correspondence: S. Ballegaard, Nordic Heart Center Ltd., Skodsborg Strandvej 198, 2942 Skodsborg, Denmark.

## Abstract

**Background:** People with ischemic heart disease (IHD) remain at high risk of recurrent major cardiovascular events despite contemporary therapy. Over two decades, a translational research program has evaluated pressure pain sensitivity (PPS) as a non-invasive marker of central autonomic dysfunction and a mutual risk phenotype in IHD and type 2 diabetes. A PPS-guided non-pharmacological intervention has been shown to substantially reduce five-year all-cause mortality in IHD.

**Methods:** In a randomized controlled trial, 213 adults with stable IHD and elevated PPS, suggesting ANSD, were allocated to PPS-guided intervention (n=106) or control (n=107). The active group received three months of structured education (daily PPS self-measurement, cutaneous sensory nerve stimulation, supportive mental and physical exercises, telemedical feedback) followed by self-directed continuation. Controls received a booklet on general stress-management. The primary endpoint for this prespecified secondary analysis was a composite of eight major cardiovascular events.

**Results:** Over 5 years, at least one major adverse cardiovascular event occurred in 19.8% of the PPS-guided group versus 43.8% of controls (odds ratio 0.32, 95% CI 0.17–0.62, P=0.0003). Incidence rates were directionally in favor of active intervention across all event categories (P=0.004).

**Conclusions:** A brief PPS-guided non-pharmacological intervention, followed by self-directed continuation, was associated with a marked long-term reduction in major adverse cardiovascular events, complementing previously reported large reductions in all-cause mortality in the same cohort. Within the context of a multi-decade PPS research program, these findings support PPS-guided care as a low-resource autonomic intervention ready for pragmatic scale-up testing as an adjunct to cardiometabolic care.

## 1. Introduction

Over more than a decade decades, a coordinated translational research program has investigated periosteal/pressure pain sensitivity (PPS) as a non-invasive marker of central autonomic nervous system dysfunction and as a guide for a structured, non-pharmacological intervention across healthy people, people with ischemic heart disease (IHD), type 2 diabetes (T2D), type 1 diabetes (T1D) and depression. Across experimental studies, observational cohorts, and randomized controlled trials, PPS-guided intervention has been associated with improvements in autonomic regulation, cardiovascular risk factors, mental health, glycemic control, and long-term survival, suggesting that PPS reflects a common stress–autonomic phenotype that can be modified by targeted intervention (1–4). Within this context, the present analysis of major adverse cardiovascular events in IHD represents a pivotal efficacy step in a multi-decade PPS program, providing clinical outcome evidence to justify multisite pragmatic real-life trials, including in resource-constrained health-care settings where low-cost non-pharmacological strategies are urgently needed (1, 2, 5).

Ischemic heart disease (IHD) remains the leading cause of morbidity and mortality globally, despite substantial advances in pharmacological and interventional management. While contemporary medical therapies effectively address hemodynamic parameters and atherosclerotic burden, the persistent burden of recurrent cardiovascular events suggests that important pathophysiological mechanisms remain inadequately targeted. Among these mechanisms, the role of autonomic nervous system (ANS) dysregulation in precipitating acute cardiac events has received increased recognition in recent years (6, 7). Thus, emerging evidence suggests that targeting ANS dysfunction (ANSD) may offer complementary benefits beyond conventional interventions (6, 7). The theoretical basis for ANS modification as a cardiovascular protective strategy rest on established evidence linking increased sympathetic activity and/or reduced parasympathetic activity to cardiovascular disease risk and mortality (8–10), and showing that abnormalities in heart rate variability predict increased coronary heart disease risk and mortality from multiple causes (7). As such, autonomic imbalance, characterized by sympathetic hyperactivity and/or parasympathetic hypoactivity, represents a common pathway to increased cardiovascular morbidity and mortality (8).

Periosteal pressure sensitivity (PPS), measured as the minimum force required to elicit pain sensation at the sternum, has been proposed as a non-invasive surrogate marker of central ANS function (1). Elevated PPS has been associated with abnormal pain processing, autonomic imbalance, and an exaggerated stress response phenotype, conditions theoretically linked to increased cardiovascular vulnerability (1). The mechanistic basis underlying this association involves dysregulation of descending pain inhibitory pathways that share neurobiological substrates with autonomic control systems, particularly those regulated by orexin-mediated arousal and the polyvagal system governing cardioprotective parasympathetic tone (6). Recent evidence demonstrates that vagal modulation of stress responses, physical exercise, and psychosocial factors substantially influence resting heart rate and autonomic tone regulation, providing mechanistic support for the PPS–ANS hypothesis (1).

The present trial forms part of a research program initiated when prospective observational studies in a private outpatient clinic indicated that a conceptual approach adding PPS-guided non-pharmacological intervention to conventional care was associated with substantial reductions in all-cause mortality and in the use of both general and cardiac-specific health-care services in people with IHD (2). The intervention used was a structured PPS-guided non-pharmacological intervention that combined daily PPS self-assessment and feedback with cutaneous sensory nerve stimulation and supportive mental and physical exercises, aiming to normalize elevated PPS and thereby modulate central stress and autonomic regulation (2). The nature of the intervention has remained basically unchanged, apart from incorporating incremental user-feedback and the development of a medical device to quantify PPS (2).

These unexpectedly large effects raised the question of whether the observations were coincidental or reflected a reproducible, biologically plausible mechanism. A stepwise sequence of mechanistic, observational, and randomized controlled trials was then conducted, to (i) clarify the physiological meaning of the PPS measure, (ii) determine whether PPS-guided intervention could be replicated under RCT conditions, and (iii) explore whether this approach might provide a beneficial adjunct in health care (2–4). Early studies in healthy individuals identified elevated PPS as a correlate of persistent stress, adverse cardiovascular risk profiles, and increased symptom burden (2, 3). Subsequent randomized and mechanistic trials in IHD and T2D, with and without depression, demonstrated that elevated PPS was associated with altered central autonomic regulation, and that PPS-guided intervention targeting PPS reduction correlated with improvements in autonomic function, cardiovascular risk factors, mental health (i.e.,depression score), and glycemic control as an indication of mutual pathophysiological mechanisms (2–4).

In previous work, a structured PPS-guided non-pharmacological intervention was developed that combines daily PPS self-measurement and feedback with cutaneous sensory nerve stimulation and supportive mental and physical exercises, aiming to normalize elevated PPS and thereby modulate central stress and autonomic regulation (2). In a cohort of people with stable IHD, using a RCT design, PPS-guided intervention was associated with a substantial 82% reduction in five-year all-cause mortality compared with a passive intervention control group, and a subsequent meta-analysis of three consecutive studies (one RCT, and two prospective observational cohort studies) in people with cardiovascular disease indicated approximately 60% lower all-cause mortality than expected from the Danish background population (2). Within this incremental framework, the current cardiovascular-event analysis represents a crucial next step by examining whether the observed all-cause mortality benefit associated with PPS reduction is paralleled by a reduction in a broad spectrum of major cardiovascular events plausibly linked to reversal of central autonomic nervous system dysfunction (2).

In the present secondary analysis of data from the same RCT in a cohort of people with IHD (2), the selection of eight predefined, unique major adverse cardiovascular events was pragmatic and the major adverse cardiovascular events(MACE) III composite endpoint was included as an accepted composite endpoint. The components were restricted to events already available in the Danish Herlev–Østerbro trials (5) and chosen to fit the aim of capturing a broad spectrum of autonomically related cardiovascular outcomes. Because autonomic dysfunction is implicated not only in cardiac death but also in myocardial infarction, stroke, atrial fibrillation, heart failure, and peripheral arterial disease, we hypothesized that PPS would be associated with a broad spectrum of serious cardiovascular events rather than with events strictly attributable to an atherosclerotic burden alone (1, 2, 5, 8). On this background, we tested the hypothesis that a PPS-guided non-pharmacological intervention, aiming to reduce an elevated PPS as a sign of reversal of central ANSD, would reduce major adverse cardiovascular events in people with established IHD, thereby helping to explain the substantial mortality reduction previously observed (2), and informing the rationale for future pragmatic real-life scale-up trials.

## 2. Materials and Methods

### 2.1. Ethics

The study was approved by the local ethics committee (The Regional Ethics Committee of the Copenhagen Region, Kongensvænge 2, DK-3400 Hillerød, www.regionh.dk/vek, identifier H-4-2010-135, and amendment 31962) and by the Danish Data Protection Agency (identifier 2011-41-7022), registered on www.clinicaltrials.gov (identifier NCT01513824). All participants gave their written informed consent after oral and written information about the study. The study was performed according to the declaration of Helsinki. AI has been used for language correction.

### 2.2. Design and Participants

This secondary analysis utilized data from a prospective, parallel-group randomized controlled trial of 213 people with documented IHD (prior myocardial infarction, percutaneous coronary intervention, or coronary artery bypass grafting). Participants were recruited from cardiac rehabilitation centers and cardiology outpatient clinics, and all participants provided written informed consent. The practical conduction of the study took place at Herlev-Gentofte University Hospital, Medical department. The study protocol was approved by the Danish Data Protection Agency (identifier 2011-41-7022), and all procedures conformed to the Declaration of Helsinki principles. The calendar years of enrollment into this study included 2011 and 2012 for 80 and 26 patients of the active intervention group and for 79 and 28 patients of the passive intervention group, respectively. The period of study ended 31 December 2017.

Inclusion criteria encompassed adults aged 40–75 years with angiographically or functionally documented evidence of coronary artery disease. Exclusion criteria included acute coronary syndrome within the preceding three months, left ventricular ejection fraction <30%, severe valvular disease, severe renal or hepatic impairment, or inability to comply with study procedures. For additional details, see reference (2).

Although the present study was conceived and conducted as a traditional randomized controlled trial, several design choices were made with future implementation and scalability in mind. First, participants were recruited from routine cardiac rehabilitation centers and cardiology outpatient clinics, mirroring typical secondary-prevention settings rather than highly selected tertiary referral cohorts. Second, the PPS-guided intervention was deliberately limited to a three-month structured education phase, after which participants continued the intervention self-directed without scheduled provider contact, reflecting a pragmatic model that could be integrated into existing services with minimal additional resource use. Third, the use of daily PPS self-measurement and telemedical feedback was intended to explore a delivery mode that could, in principle, support future low-cost, remotely supervised implementation, including in resource-constrained health-care systems (2, 5).

### 2.3. Randomization and Intervention

Participants were randomized 1:1 to either the active or passive intervention using computer-generated allocation sequences with stratification by age and prior revascularization status. Group allocation was concealed until after baseline assessment.

#### Active Intervention

Participants attended three months of unique, trademark-protected structured education sessions delivered by trained health professionals, during which they received detailed instruction on techniques specifically designed to normalize elevated PPS measurements. These techniques incorporated: 1) daily PPS self-measurement combined with cognitive reflection on daily stress levels, 2) daily cutaneous sensory nerve stimulation at specific anatomical locations with the aim to reduce an elevated PPS and subsequently maintain a non-elevated PPS, 3) free-of-choice mental and physical exercises supporting this aim. 4) daily recording of the PPS-measure allowing telemedical support in case the PPS-measure did not decline or was missing. This was performed during the initial educational period of 3 months. We encouraged participants to conduct nerve stimulation ad hoc in case of angina pectoris. We instructed a spouse in daily cutaneous nerve stimulation at the back of the chest of the subject as a preventive measure, including ad hoc measures in cases of present angina. Critically, formal contact between participants and the research team was eliminated after the initial three-month education period; participants were instructed that the intervention had previously demonstrated efficacy in healthy populations for reducing elevated PPS and were encouraged to continue self-directed implementation based on the provided education and materials. The intervention has previously been described in detail (2).

#### Passive Intervention

Participants in the passive intervention group received detailed information explaining that elevated PPS represented a measurable physiological sign of persistent stress and carried potential negative prognostic implications. They received a comprehensive educational booklet containing general stress management recommendations, relaxation techniques, and information about stress-related health effects. No formal guidance regarding specific intervention strategies was provided.

### 2.4. Outcome Measures and Definitions

The primary outcome for this analysis was the composite incidence of 8 unique major adverse cardiovascular events, defined as the occurrence of any of the following: cardiovascular death (sudden cardiac death or death attributable to acute myocardial infarction, heart failure, or arrhythmia), non-cardiac death, non-fatal myocardial infarction, non-fatal stroke, new-onset atrial fibrillation, new-onset heart failure requiring medical treatment, new peripheral arterial disease, or coronary artery bypass grafting (CABG). Individual event categories were also examined separately. A secondary endpoint was MACE III, defined as non-fatal myocardial infarction, non-fatal stroke, and cardiovascular death.

These eight components were selected a priori in collaboration with an experienced cardiovascular statistician, based on the events that could be reliably ascertained from the Danish nationwide registers (doi:10.1136/bmjopen-2016-012832) without additional adjudication work. This choice ensured complete and unbiased follow-up for all participants while still covering a broad spectrum of serious cardiovascular outcomes.

Our aim was to test whether the previously observed reduction in all-cause mortality associated with lower pressure pain sensitivity (PPS) and PPS-guided intervention, interpreted as reflecting improved central autonomic nervous system (ANS) regulation, extends beyond mortality to encompass a wider range of ANS-related cardiovascular events. The composite endpoint was therefore designed to capture both fatal and non-fatal manifestations of cardiovascular disease plausibly linked to autonomic dysfunction, rather than to isolate any single disease entity.

Cardiovascular events were identified through the unique 10-digit Danish civil registration number (CPR), which tracks all registered individuals in the Danish healthcare system. This identifier enabled determination of whether each participant experienced any of the 8 distinct cardiovascular events during the follow-up period through linkage with national hospital discharge registries and vital statistics databases. Due to local data regulations, we presented numbers of clinical events of less than or equal to three in the statistical analysis as “three or fewer events”.

### 2.6. Blinding

The professional instructor measuring PPS was blinded regarding all results of online questionnaires as well as to the results of the randomization. Further the PPS device was designed in away making the measure non-visible before the end of each measurement for both instructor and patient. The patients were instructed before randomization not to reveal the result of the randomization to the research personnel performing the follow up investigations at three months. Statistical analysis was performed prior to the unveiling of the randomization codes.

### 2.7. Sample size calculation

Sample size calculation was conducted in association with the original RCT, in which the focus was effect on depression score (2).

### 2.8. Statistics

The primary analysis compared the cumulative incidence of the 8 unique cardiovascular events between active and passive groups using Fisher’s exact test and calculation of odds ratios and 95% confidence intervals. Odds ratios were calculated from 2×2 contingency tables. When zero cell counts occurred, a continuity correction was applied by adding 1 to all cells to enable estimation of the odds ratio. Individual event types were compared using similar methods. All analyses followed the intention-to-treat principle, with participants remaining in their assigned groups regardless of adherence to intervention components. The choice of Fisher’s exact test was the predefined selected method, as this method allows to record the full burden from cardiovascular events for the individual participant, and as such providing the ability for distinguishing an initial minor event (e.g., new atrial fibrillation from a later more serious event, e.g., non-fatal stroke or death). The rationale for this choice is illustrated in Table 3. However, a Kaplan Meyer analysis is a common method to elucidate event rates, and we conducted this analysis for MACE III, as shown in Figure 3.

By chance the prevalence of diabetes was significantly higher in the active group. For that reason, we additionally performed a secondary exploratory stratified analysis using the Mantel–Haenszel method to obtain odds ratios adjusted for baseline diabetes status. Because only a minority of participants had diabetes at baseline, the stratified Mantel–Haenszel analyses adjusting for diabetes were based on relatively small numbers within the diabetes stratum. These adjusted odds ratios were therefore considered exploratory robustness analyses rather than primary estimates of treatment effects within diabetes subgroups and are reported as supplementary to the pre-specified Fisher-based analyses.

Two-sided statistical analyses are presented, only although the tested hypothesis is strictly one-directional

## 3. Results

### 3.1. Participant Characteristics and Follow-up

Of 213 randomized participants, 106 were allocated to active intervention and 107 to passive intervention. Follow-up data were available for 104 (99%) of the active group and 105 (98%) of the passive group (Figure 1). Baseline demographic, clinical, and psychometric characteristics were well balanced between groups, with no significant differences in age (mean 62 years), gender distribution (73% male), prior myocardial infarction prevalence (64%), or revascularization history (Table 1).

**Figure 1.**
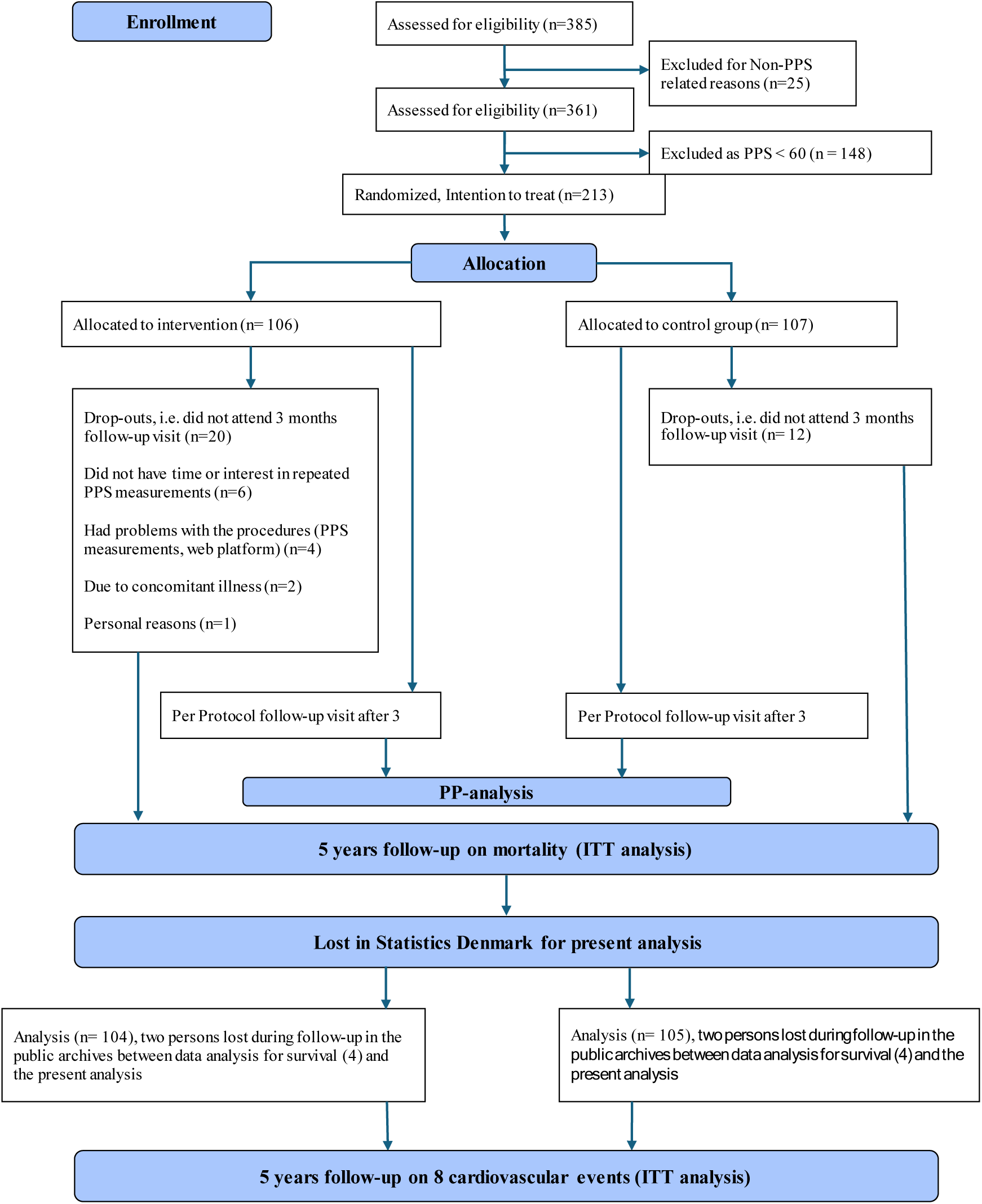
Consort diagram.

**Table 1.** Distribution of baseline factors according to RCT treatment groups.

|  | Full sample | Active group | Passive group | p-value |
| --- | --- | --- | --- | --- |
| Number of participants (n) | 213 | 106 | 107 |  |
| Male (n, %) | 156 (73 %) | 78 (74 %) | 78 (73 %) | NS* |
| Age in years (mean, SD) | 62 (8.1) | 62 (8.1) | 62 (8.2) | NS |
| *Psychometrics* |  |  |  |  |
| MDI (mean, SD) | 8.9 (7.4) | 8.4 (7.7) | 9.4 (7.0) | NS |
| WHO-5 (mean, SD) | 65 (19) | 67 (19) | 63 (19) | NS |
| PPS (mean, SD) | 81 (13) | 81 (13) | 81 (13) | NS |
| SF-36 PCS (mean, SD) | 48 (8.4) | 48 (9.1) | 48 (7.6) | NS |
| SF-36 MCS (mean, SD) | 52 (9.3) | 53 (9.3) | 52 (9.3) | NS |
| CSS (mean, SD) | 9.7 (7.1) | 9.2 (6.5) | 10 (7.6) | NS |
| *Social status* |  |  |  |  |
| Married or cohabitating (n, %) | 175 (82 %) | 83 (78 %) | 92 (86 %) | NS |
| Have children (n, %) | 190 (92 %) | 97 (91 %) | 96 (90 %) | NS |
| *Employment status* |  |  |  |  |
| Employed (n, %) | 106 (50 %) | 54 (51 %) | 52 (49 %) | NS |
| Unemployed (n, %) | 4 (2 %) | 3 (3 %) | 1 (1 %) | NS |
| Retired (n, %) | 92 (47 %) | 46 (44 %) | 52 (48 %) | NS |
| *Cardiac variables* |  |  |  |  |
| Self-reported time (years) since first AMI (mean, SD) | 7.5 (5.8) | 8.2 (6.5) | 6.8 (5.0) | NS |
| Previous AMI (n, %) | 136 (64 %) | 69 (65 %) | 67 (63 %) | NS |
| Treated with PCI (n, %) | 147 (69 %) | 73 (69 %) | 74 (69 %) | NS |
| Treated with CABG (n, %) | 52 (24 %) | 27 (25 %) | 25 (23 %) | NS |
| In-hospital days during last 12 months<br>before inclusion due to cardiac disease (n,<br>(%), mean number of days) | 46 (22%)<br>6 days | 18 (17%)<br>6 days | 28 (26%)<br>7 days | NS |
| Visits Cardiac Outpatient Clinic during last<br>12 months before inclusion (n (%), mean<br>number of visits) | 68 (32%),<br>3 visits | 27 (25%)<br>4 visits | 41 (39%)<br>3 visits | NS |
| Visits to Cardiologist during last 12 months<br>before inclusion (n (%), mean number of<br>visits) | 29 (16%)<br>2 visits | 13 (12%)<br>2 visits | 16 (15%)<br>2 visits | NS |
| Visits to General Practitioner during last 12<br>months before inclusion for cardiac disease<br>(n (%), mean number of visits) | 90 (47%)<br>3 visits | 46 (43%)<br>3 visits | 44 (42%)<br>2 visits | NS |
| CCS Angina Pectoris Class I | 55 (26%) | 26 (25%) | 29 (27%) | NS |
| CCS Angina Pectoris Class II | 25 (12%) | 14 (14%) | 11 (10%) | NS |
| CCS Angina Pectoris Class III | 5 (2%) | 1 (1%) | 4 (4%) | NS |
| CCS Angina Pectoris Class IV | 2 (1%) | 1 (1%) | 1 (1%) | NS |
| Chest Pain at rest | 71 (34%) | 39 (38%) | 32 (30%) | NS |
| Resting pulse (mean, SD) | 61 (11) | 61 (11) | 60 (11) | NS |
| MAP (mean, SD) | 98 (10) | 98 (9.7) | 97 (11) | NS |
| *Cardiac risk factors* |  |  |  |  |
| BMI (mean, SD) | 27.6 (4.3) | 27.8 (4.3) | 27.4 (4.4) | NS |
| Triglyceride (mean, SD) | 1.5 (0.9) | 1.4 (0.7) | 1.5 (1.0) | NS |
| Current smoker (n, %) | 22 (10 %) | 9 (9 %) | 13 (12 %) | NS |
| Heart Rate ≥ 70 beats/minute (n, %) | 35 (16%) | 20 (19 %) | 15 (14%) | NS |
| *Self-reported co-morbidity* |  |  |  |  |
| Heart failure (n, %) | 72 (34 %) | 29 (27 %) | 43 (40 %) | NS |
| Chronic obstructive lung disease (n, %) | 13 (6 %) | 5 (5 %) | 8 (8 %) | NS |
| Diabetes (n, %) | 28 (13 %) | 20 (19 %) | 8 (8 %) | P = 0.015 |
| Previous cerebral insults (n, %) | 15 (7 %) | 7 (7 %) | 8 (8 %) | NS |
| Have been treated for depression (n, %) | 32 (15 %) | 12 (11 %) | 20 (19 %) | NS |
| Elevated Depressions score (i.e., MDI score ≥ 15, indication incipient depression) | 41 (19%) | 21 (20%) | 20 (19%) | NS |
| *Medication* |  |  |  |  |
| Beta-blockers (n, %) | 125 (60 %) | 65 (61 %) | 60 (57 %) | NS |
| Cholesterol-lowering medication (n, %) | 188 (90 %) | 94 (89 %) | 94 (88 %) | NS |
| Calcium antagonists (n, %) | 47 (23 %) | 26 (25 %) | 21 (20 %) | NS |
| Angiotensin-II antagonist and/or ACE inhibitors (n, %) | 115 (55 %) | 56 (53 %) | 59 (55 %) | NS |
| Diuretics (thiazide or furosemide) (n, %) | 74 (36 %) | 40 (39 %) | 34 (33 %) | NS |
| Anti-depressive medication (n, %) | 12 (6 %) | 4 (4 %) | 8 (8 %) | NS |

\*NS: $p > 0.05$ between active group and passive; BMI: Body mass Index; MDI: Major Depressive Inventory; WHO-5: quality of life questionnaire. SF-36 PCS: Short form self-report health questionnaire Physical component summary; SF-36 MCS: Short form self-report health questionnaire component summary; CSS: Clinical stress symptom score questionnaire; AMI: acute myocardial
infarction; MAP: mean arterial blood pressure; CCS Angina Class: Canadian Cardiovascular Society angina score; BMI: body mass index.

### 3.2. Primary Outcome: Major Adverse Cardiovascular Events

Over the 5-year follow-up period, major adverse cardiovascular events occurred in 21 participants (19.8%) in the active group compared with 46 participants (43.8%) in the passive group. This difference corresponds to an odds ratio of 0.32 (95% CI 0.17–0.62, P=0.0003), representing a 68% relative risk reduction in the composite of 8 distinct event incidences for the active intervention group. In a secondary, stratified Mantel–Haenszel analysis adjusting for baseline diabetes status, the odds ratio was 0.52 (95% CI 0.25–1.07; p = 0.07).

Notably, all eight events showed numerically lower frequencies in the active group than in the passive group (P = 1/2^8^ = 1/256 = 0.004). The protective effect of the active intervention was distributed across multiple event categories rather than concentrated in a single endpoint. Table 2 displays the detailed breakdown of the eight cardiovascular event categories. Table 3 shows the number of participants with their number of cardiovascular events. The odds ratio for having no event during the five years of observation, and when comparing active and control group is 1.9 (95 % CI :1.0 −3.6; P=0.05). Notably, only one participant in the active group experienced more than one cardiovascular event during follow-up, compared with seven participants in the passive group; although the numbers are too small for formal statistical testing, this pattern is directionally consistent with the overall reduction in event incidence in the active group.

**Table 2.** Incidence of Eight Major Adverse Cardiovascular Events by Group.

| **Event Category** | **Active**<br>**(n=104)** | **Passive**<br>**(n=105)** | **Odds**<br>**Ratio** | **P-Value** |
| --- | --- | --- | --- | --- |
| Cardiovascular death | ≤ 3* | ≤ 3* | 0.14 | 0.25 |
| Non-cardiovascular death | ≤ 3* | 5 | 0.39 | 0.45 |
| Non-fatal myocardial infarction | 6 | 7 | 0.86 | 1.00 |
| Non-fatal stroke | ≤ 3* | 7 | 0.14 | 0.07 |
| New-onset atrial fibrillation | ≤ 3* | 7 | 0.42 | 0.33 |
| New-onset heart failure | 5 | 7 | 0.71 | 0.77 |
| New peripheral arterial disease | 4 | 7 | 0.56 | 0.54 |
| Coronary artery bypass grafting | ≤ 3* | ≤ 3* | 0.14 | 0.25 |
| **Total of 8 individual**<br>**cardiovascular events** | **21** | **46** | **0.32** | **0.0003** |

\*Due to local data regulations, we present numbers of clinical events of less than or equal to three in as “three or fewer events”. However, in the statistical analysis the actual numbers of events are used.

**Table 3.**
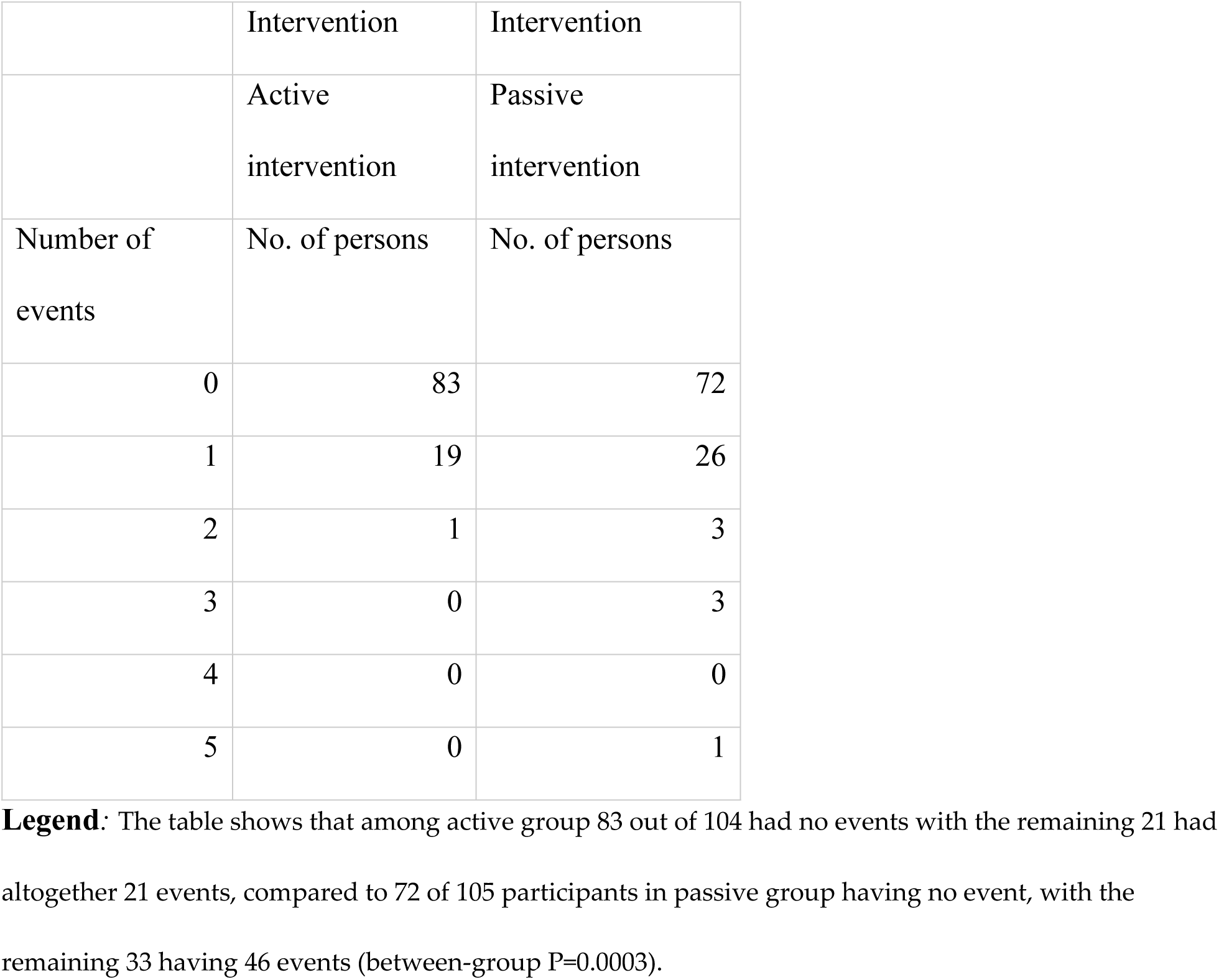
Incidence of Eight Major Adverse Cardiovascular Events by the individual participant.

### 3.3. MACE III Analysis

A secondary composite endpoint combining only the three most clinically significant events—non-fatal myocardial infarction, non-fatal stroke, and cardiovascular death—yielded similarly results. This more restrictive composite occurred in 7 participants (6.7%) in the active group versus 17 participants (16.2%) in the passive group (odds ratio 0.37, 95% CI 0.15–0.93, P=0.049) (Figure 2).

**Figure 2.**
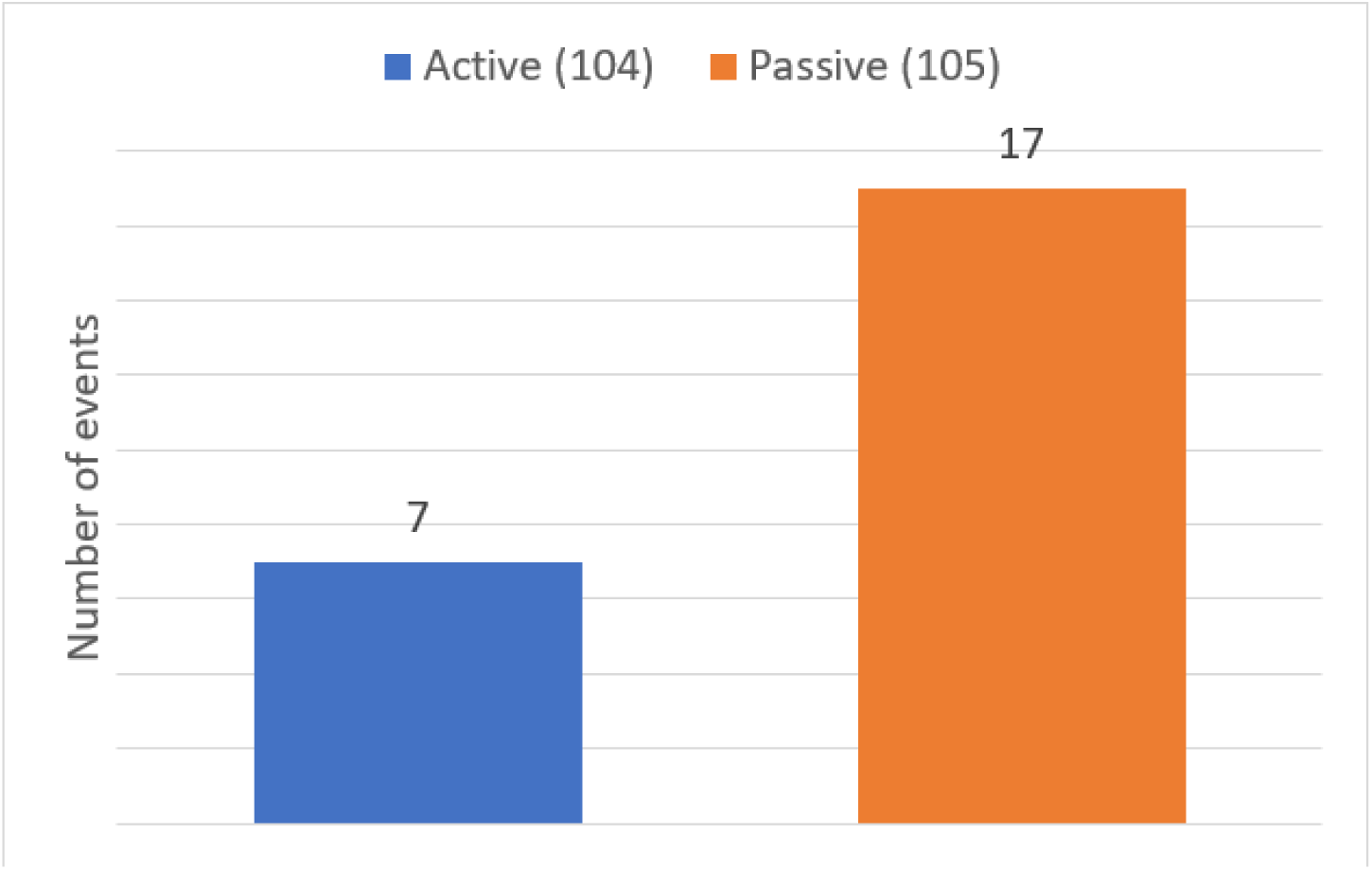
Comparison of MACE III between active and control group. **Legend:** Comparison of incidence of MACE III (non-fatal myocardial infarction, non-fatal stroke, and cardiovascular death) between active and passive groups (P = 0.049).

The secondary diabetes-adjusted Mantel–Haenszel analysis yielded a similar odds ratio of 0.42 (95% CI 0.18–0.99; p = 0.043). Figure 3 shows the Kaplan Meyer analysis, adjusted for diabetes.

**Figure 3.**
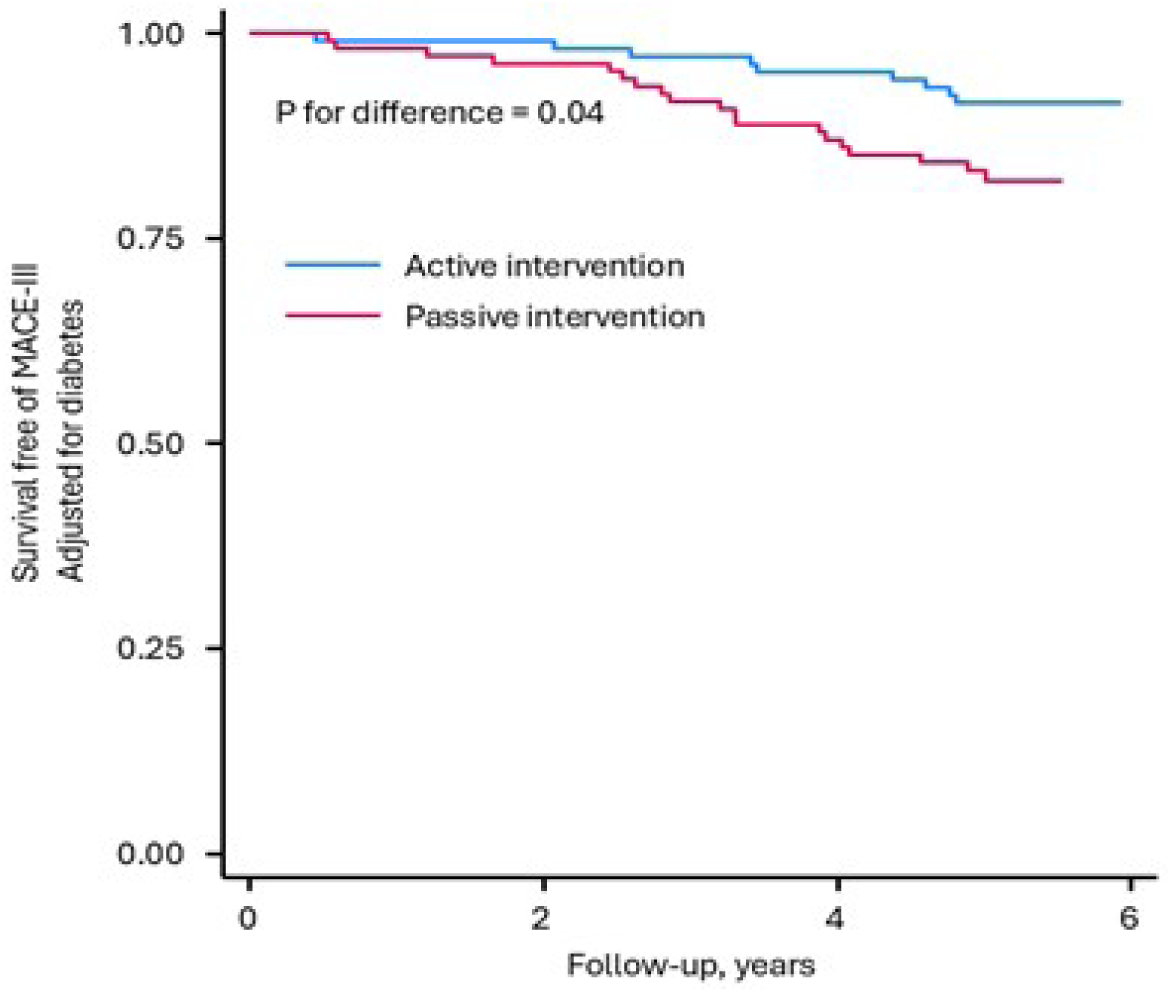
Comparison of Kaplan Meyer analysis regarding MACE III for active and control group. **Legend:** Comparison of Kaplan Meyer analysis regarding MACE III (non-fatal myocardial infarction, non-fatal stroke, and cardiovascular death) for active and control group adjusted for diabetes (P=0.043).

### 3.4 Safety

As no contact to participants were included after the initial 3-months of education, no formal registration of adverse effects was registration for the device. However, no participant contacted the research site with any such observations. As no pharmacological treatment was involved, adverse events related to the intervention were not monitored systematically.

## 4. Discussion

Ischemic heart disease (IHD) continues to impose a major burden of recurrent cardiovascular events despite contemporary evidence-based pharmacotherapy and revascularization, underscoring the need for adjunctive, scalable prevention strategies that can be integrated into routine care. In this randomized trial of people with stable IHD and elevated pressure pain sensitivity (PPS), a brief, three-month PPS-guided non-pharmacological intervention, followed by self-directed continuation without further provider contact, was associated with a substantial reduction in major cardiovascular events over five years compared with a passive stress-management control. These findings extend previous observations of markedly reduced all-cause mortality in the same cohort and, when viewed together with earlier mechanistic and clinical studies across healthy people, people with IHD, T2D, T1D, and depression, support conceptualization of PPS-guided intervention as a mature autonomic platform that is now ready for pragmatic real-life scale-up testing, particularly in resource-constrained settings where low-cost, non-pharmacological options are especially valuable.

### 4.1. Summary of Findings

In this randomized analysis of people with stable IHD and elevated PPS, a three-month education into a PPS-guided non-pharmacological intervention was associated with a 68% relative reduction in the five-year composite incidence of eight major cardiovascular events and a 63% relative reduction in MACE III events, compared with a passive intervention. These findings extend our previous report of an 82% reduction in all-cause mortality in the same cohort (2). The present results ad converging evidence that PPS-guided intervention not only reduces all-cause mortality but also lowers the incidence of a broad spectrum of major cardiovascular events, supporting the hypothesis that targeting central autonomic dysfunction can favorably modify cardiovascular vulnerability in this population.

The primary analyses in this study were based on Fisher’s exact test, as specified a priori, to accommodate the modest sample size and small cell counts for several event categories. Given the higher prevalence of diabetes at baseline in the active group, we additionally performed stratified Mantel–Haenszel analyses adjusting for diabetes status. These secondary analyses yielded diabetes-adjusted odds ratios of 0.52 for the 8-event composite and 0.42 for MACE III, with the latter remaining statistically significant. Thus, although adjustment for diabetes reduced the statistical precision for the 8-event composite and led to loss of conventional significance, which is somewhat counter-intuitive given diabetes prevalence was higher in the active group, the estimated effect sizes remained large and directionally consistent. This pattern suggests that the observed reductions in cardiovascular events cannot be explained by the baseline imbalance in diabetes alone, while also highlighting the limited power for adjusted analyses in this relatively small trial.

### 4.2. Modulation of Sensitivity to Periosteal Pressure and Other Mechanistic Considerations

We define neuromodulation as mechanisms of chemical, electrical, or mechanical pain induction, as well as modulation of autonomic sympathetic activity, that generate afferent impulses to the brain (11). Within this framework, different methods of sensory nerve stimulation may nevertheless converge on similar central autonomic nervous system (ANS) structures, with the specific functional outcome depending on the characteristics of the stimulation. Established neuromodulation techniques include vagal and sacral nerve stimulation (12–14), spinal cord stimulation (15, 16), and non-noxious cutaneous sensory nerve stimulation (17, 18). Non-noxious sensory nerve stimulation has been reported to reduce symptoms of diabetic neuropathy (15) and to reduce the number of angina pectoris attacks in people with IHD (16). These effects resemble the well-known autonomic reflex arc regulating pain sensation through diffuse noxious inhibitory control and related mechanisms that modulate efferent responses (19, 20).

Non-noxious cutaneous sensory stimulation is also a well-established treatment modality in pre-term newborn infants, where skin-to-skin contact has been associated with reduced stress and improved survival (17, 21). A similar principle is incorporated into the present intervention, in which non-noxious cutaneous stimulation by moderate pressure at the back is applied twice daily, typically by a spouse or partner, as part of the PPS-guided program. Within this broader neuromodulation context, the present findings are consistent with the concept that autonomic nervous system dysregulation is a central pathway linking psychosocial stress to acute cardiovascular events (11).

Elevated PPS is hypothesized to reflect central sensitization of nociceptive pathways together with abnormal autonomic regulation, including sympathetic hyperactivity and reduced parasympathetic buffering. Such imbalance can promote arrhythmia and hemodynamic and myocardial electrical instability via brainstem circuits, including the locus coeruleus and dorsal raphe nucleus, that govern both pain modulation and sympathetic outflow (12). In an experimental study in the same group of people with IHD as the present study, the three-month educational period implementing the PPS-guided intervention was associated with a reduction in PPS and a concomitant increase in the heart rate variability response to tilting. Furthermore, the magnitude of PPS reduction was directly associated with improved baroreflex responses to tilting, including increased systolic blood pressure reactivity (13). These observations support the interpretation, that PPS-guided intervention can shift autonomic regulation towards a more resilient sympathetic-vagal balance.

Other recent studies suggest that the effects of reducing an elevated PPS are mediated by central autonomic pathways. Thus, two independent RCTs in IHD and T2D, respectively, found that elevated PPS was associated with higher depression scores and altered autonomic regulation of mood (submitted). In these studies, reductions in elevated PPS during the PPS-guided intervention correlated to improvements in autonomic regulation of mood as well as in depression scores, indicating a link between PPS modulation, ANS function, and mood regulation (submitted). In people with T1D, an RCT showed that reduction of an elevated PPS was associated with a concomitant reduction in reduction in the total number of elevated individual cardiovascular risk factors (submitted). In healthy women, a small randomized cross-over study found that salivary orexin, which is accepted as a key regulator of ANS function, was positively associated with PPS, consistent with a potential role of orexin-mediated arousal pathways in PPS and autonomic regulation (submitted).

Taken together, these observations support the view, that reductions in elevated PPS are accompanied by a shift toward a more balanced sympathetic and parasympathetic tone, likely mediated by central autonomic mechanisms, and possibly involving orexin-sensitive pathways in the lateral hypothalamus. The three-month intensive education phase may induce durable changes in autonomic regulation as well as in stress-coping behavior through neural plasticity. In line with this, we have previously observed increased empowerment in people with T2D receiving the same intervention, suggesting that PPS-guided education can strengthen self-management and engagement with lifestyle changes (22). The subsequent self-directed, year-long period in the present trial may allow these behavioral and autonomic adaptations to consolidate, providing a plausible explanation for the sustained protection observed over five years. In the present study we did not have 5-year data regarding the PPS-measurement, which excludes the possibility to elucidate a direct mechanistic link between reduction of PPS and reduction of mortality and cardiovascular event rate, as this scope was outside the aim of the study. The present outcome invites future studies to elucidate this important mechanistic question.

### 4.3. Relation to Previous Evidence

The accumulated findings from early observations studies and subsequent randomized controlled trials showing improvements in cardiovascular risk factors, symptoms, and quality of life in both healthy individuals, people with IHD, and people with T2D (2, 4). the present data suggest that the PPS-guided intervention platform targeting ANSD can exert beneficial effects both at the level of cardiovascular risk profiles, and at the level of clinical events and survival in IHD.

These findings substantially expand upon the large mortality reduction reported in our previous analysis of this cohort as well as upon the subsequent meta-analysis showing a marked reduction in all-cause mortality when compared to the Danish general population (2). As such the present findings demonstrate a parallel reduction in a composite measure of eight distinct major cardiovascular events in the same RTC population, and each event with plausible links to ANS regulation.

### 4.4. Clinical and Public Health Implications

Integration of PPS assessment and targeted intervention into routine post-myocardial infarction and post-revascularization care could potentially enhance the protective benefits achievable through current standard practices (23). The non-pharmacological nature of the intervention, the absence of observed treatment-related side effects or complications, its simplicity, and the limited need for ongoing provider contact make this approach potentially attractive for diverse healthcare settings, including resource-constrained environments.

Along this line, future studies in IHD should combine advanced autonomic and neurobiological mechanistic assessment with pragmatic trial designs that explicitly test different delivery and scale-up models, including cluster-randomized trials in routine care and in low-resource settings, and incorporate formal cost-effectiveness and cost-utility analyses.

### 4.5. Study Strengths and Limitations

The study has several strengths, including the collection of data from a public register: the Herlev-Østerbro survey (5), It is also a strength that our endpoint strategy is of pragmatic nature: By restricting the composite to events already available in the Herlev–Østerbro infrastructure (5), we achieved virtually complete and unbiased follow-up without post-hoc endpoint selection. At the same time, the chosen components represent a clinically and mechanistically coherent group of serious cardiovascular outcomes with established or plausible links to autonomic nervous system dysfunction.

In contrast to the general challenge of low adherence to traditional non-pharmacological cardiac rehabilitation programs (23, 24), the special features of the present active intervention program aimed at personal adherence and compliance. As such, a persistently high compliance rate above 80% to the intervention program during follow-up periods of 3-6 months was found in 5 consecutive studies across a variety of population groups including healthy persons, women with breast cancer, people with T1D, T2D and IHD (2, 22). We considered this evidence as a strength with respect to the daily adherence to the present cardiac rehabilitation program, although the adherence probably will drop somewhat during time. Unfortunately, we have no data upon this particular issue.

In addition, we consider the use of a RCT design and using ITT for the statistical analyses as additional strengths of the study.

The observed 68% relative reduction in major event rate in the active group of this RCT is a surprise. For new interventions within recent decades, the reduction rarely exceeded 25%, probably reflecting that each of these interventions approached one health risk factor, only. This suggests that a reduction in an elevated PPS measure may reflect a more global impact on health, affecting a broad range of health risk factors at the same time, including mood, glucose regulation, and the number of elevated risk factors for cardiovascular disease, including targeting the metabolic syndrome profile (2, 22). Such a multi-factorial approach is consistent with the established concept that simultaneous, multifactorial risk-factor control yields large reductions in cardiovascular events and mortality (25). Furthermore, this pattern aligns with recommendations by WHO to assess and manage cardiovascular disease based on total risk, emphasizing the combined burden of multiple risk factors rather than single parameters alone. The present PPS-guided intervention can therefore be viewed as a practical, real-life implementation of this WHO-endorsed multi–risk-factor approach (26).

Several limitations must be considered: Our composite endpoint of 8 unique cardiovascular events do not correspond exactly to conventional definitions by combining heterogeneous endpoints. However, this heterogeneity is intrinsic to our mechanistic question, which concerns a generalized autonomic influence on multiple vascular beds and cardiac functions rather than a single disease entity. Addressing this limitation, we did perform a more conventional MACE III analysis, which showed similar results as the 8-point analysis.

The sample size was small but never-the-less adequate for detecting the observed effect magnitude. As such, it precludes robust detection of differences in specific event categories and limits generalizability. Our findings should therefore be interpreted primary at the level of a composite endpoint, upon which all 8 event rates are in favor of active intervention.

Complete blinding of participants and treatment providers to group assignment was not feasible given the nature of the interventions. This never-the less will mimic the real-life situation executing the intervention. Furthermore, the active group received an intervention, like a “Package” with several components, and thus without the possibility to elucidate the effect of the individual components.

The follow-up period, while substantial, was finite, and questions regarding long-term durability of benefit beyond the observation window remain unanswered.

At baseline the control group has had 28 in-hospital days compared to 18 in the active group and 41 out-clinic visits compared to 27 in the active group. Although the differences are not significant, they may represent a bias favoring the active group, potentially being more affected by the underlying cardiovascular disease.

Regarding the baseline difference in prevalence of diabetes between active and control group, we performed stratified Mantel–Haenszel as secondary analysis to adjust for the higher baseline prevalence of diabetes in the active intervention group. Since only a minority of participants had diabetes at baseline, the diabetes-adjusted estimates are based on relatively small numbers within the diabetes stratum, and as such with limited precision. The trial was not powered to detect or characterize treatment effects specifically among people with diabetes, and the diabetes-adjusted odds ratios should therefore be interpreted as exploratory robustness analyses rather than definitive subgroup effects. Regarding the 8-composite event rate measure and adjusted for diabetes, it still indicates a substantial relative risk reduction but with wider confidence intervals and loss of conventional statistical significance, while regarding the MACE III the results are supporting a robust reduction in MACE III with PPS-guided intervention even after accounting for the baseline imbalance in diabetes.

A further limitation is that most of the existing evidence on PPS and PPS-guided intervention, including the present trial, originates from a single research group and a relatively narrow set of health-care systems. This concentration of evidence may raise legitimate concerns about external validity, independence, and generalizability across different clinical, organizational, and cultural contexts. However, these concerns do not diminish the internal consistency of the current findings; rather, they underscore the need for the next phase of research to focus on multi-site pragmatic trials, including cluster-randomized designs in diverse and resource-constrained settings, explicitly aimed at independent replication, contextual adaptation, and assessment of real-world scalability and equity impact.

## 5. Conclusions

In people with stable ischemic heart disease and elevated pressure pain sensitivity (PPS), a brief 3-month PPS-guided non-pharmacological intervention, followed by self-directed continuation without further provider contact, was associated with a large and durable reduction in major adverse cardiovascular events over five years compared with passive stress-management control. This cardiovascular event benefit complements previously reported 82% lower all-cause mortality in the same randomized cohort and approximately 60% lower all-cause mortality in a subsequent meta-analysis of three consecutive cardiovascular studies, indicating that PPS-guided intervention favorably influences both survival and a broad spectrum of serious autonomically related outcomes.

Viewed within the context of a multi-decade translational research program comprising mechanistic studies and randomized trials across healthy individuals and people with ischemic heart disease, type 2 diabetes, type 1 diabetes and depression, these findings support PPS-guided intervention as a clinically mature, low risk, autonomic platform with meaningful impact on autonomic regulation, cardiovascular risk factors, mood, glycemic control and hard endpoints. The present cardiovascular-event analysis therefore represents not only a confirmation of benefit in IHD, but also a pivotal step in the progression from efficacy to implementation, indicating that the next phase of research should focus on how PPS-guided care can be implemented, sustained and evaluated equitably in routine practice.

On this basis, PPS-guided intervention now warrants pragmatic multicenter real-life trials, including cluster-randomized designs, evaluating cardiometabolic and mental-health benefits alongside reach, feasibility, fidelity, cost-effectiveness and equity in diverse, including resource-constrained, health-care settings. Such studies will be essential to determine how the PPS-guided intervention platform can be integrated at scale into routine cardiac rehabilitation and chronic disease management pathways, and to assess its potential contribution to cardiometabolic health equity.

## Declarations

## Ethics approval and consent to participate

registered on www.clinicaltrials.gov (identifier NCT01513824). All participants gave their written informed consent after oral and written information about the study. The study was performed according to the declaration of Helsinki.

## Consent for publication

Not applicable, as the manuscript does not contain any individual person’s data in any form.

## Author Contributions

JF made major contributions to all aspects of this study and manuscript preparation as the Principal Investigator; SB invented the PPS device and designed the adjunct intervention, and together with JF made major contributions to this study. AS conducted statistical analysis and interpretation of results; AH and FG contributed to the planning of the conceptual aspects of the study from the very beginning, results interpretation and manuscript preparation of the present study. All authors have read and approved the final manuscript.

## Funding

This original RCT study (2) received funding from the Johan Schrøder’s Family and Business Foundation and the Lundbeck Foundation. No specific funding was received for the present 5-year follow-up study.

## Data Availability Statement

The data underlying this article will be shared upon reasonable request to the corresponding author.

## Acknowledgments

We thank the staff at the Department of Cardiology, Herlev Gentofte Hospital, Denmark, for providing access to their database on people with cardiovascular disease who underwent cardiac rehabilitation.

## Conflicts of Interest

SB invented the PPS instrument (patent numbers PA 2004-00349 and PA 2004-00550, developed the intervention (Ballegaard Heart Disease Program® and is a shareholder of the firm that owns the PPS instrument (Nordic Heart Center LLC). To avoid bias, he was not involved in patient contact, data collection, or statistical analysis and was prohibited from admittance to the research site and any contact with people during the entire 5-year observation period. His authorship does not alter the authors’ adherence to policies on sharing data and materials. The other authors have no competing interests to declare and no relation to Nordic Heart Center LLC, including employment, consultancy, patents, products in development, or marketed products.

## Symbols, abbreviations, and acronyms

ANS: Autonomic Nervous System
ANSD: Autonomic Nervous System Dysfunction
CABG: Coronary artery bypass grafting
CCS: Canadian Cardiovascular Society grading of angina pectoris.
CONSORT: Consolidated Standards of Reporting Trials
CPR: Civil registration number (Danish)
IHD: Ischemic Heart Disease
ITT: Intention-to-Treat
MACE III: Composite of non-fatal myocardial infarction, non-fatal stroke, and cardiovascular death
PPS: Periosteal Pressure Sensitivity
T1D: Type 1 Diabetes Mellitus
T2D: Type 2 Diabetes Mellitus

## Notes

### Clinical Trial

www.clinicaltrials.gov registration number: NCT01513824). Registration date: January 17th, 2012).

### Funding Statement

The author(s) received no specific funding for this work.

### Author Declarations

The study was approved by the local ethics committee (The Regional Ethics Committee of the Copenhagen Region, Kongensvænge 2, DK-3400 Hillerød, www.regionh.dk/vek, identifier H-4-2010-135, and amendment 31962) and by the Danish Data Protection Agency (identifier 2011-41-7022),

